# Efficacy and Safety of Macitentan Across Pulmonary Hypertension Phenotypes: A Systematic Review and Meta-analysis

**DOI:** 10.64898/2026.09.22.26363639

**Authors:** Saathwika Gurram, Vipin MP, Surya Teja Avula, Vishal Subramanian, Hira Aruthi Natarajan, Harshitha Byreddy, Sanjana Palakodeti

## Abstract

**Background:** Macitentan is a dual endothelin receptor antagonist used in pulmonary arterial hypertension (PAH) and has also been studied in other forms of pulmonary hypertension (PH). Evidence across PH phenotypes has been heterogeneous. This study aims to evaluate the efficacy and safety of macitentan compared with concurrent control therapies across PH phenotypes and to explore whether treatment effects differ according to PH classification.

**Methods:** PubMed, Embase, and the Cochrane database were searched from inception through June 2026. Randomized and non-randomized comparative studies of patients with PH receiving macitentan were eligible. Prespecified efficacy outcomes were 6-minute walk distance (6MWD), pulmonary vascular resistance (PVR), mean pulmonary arterial pressure (mPAP), and cardiac index; prespecified safety outcomes included mortality and adverse events. Random-effects meta-analyses were performed using mean differences (MDs) for continuous outcomes and risk ratios (RRs) for dichotomous outcomes.

**Results:** Fourteen studies met the eligibility criteria. Macitentan was associated with a greater reduction in PVR (MD, -134.50 dyn.s.cm; 95% CI, -226.10 to -42.86) and an increase in cardiac index (MD, 0.44 L/min/m; 95% CI, 0.22 to 0.67). No statistically significant difference was observed for 6MWD (MD, 5.04 m; 95% CI, -15.06 to 25.14) or mPAP (MD, -2.70 mmHg; 95% CI, -6.46 to 1.05). In pooled safety analyses, mortality was lower and risks of anemia, peripheral edema, and discontinuation due to adverse events were higher with macitentan. Heterogeneity and the small number of studies within several phenotype-specific analyses limited the precision of subgroup findings.

**Conclusions:** Macitentan-containing therapy was associated with favorable changes in selected hemodynamic outcomes but not with a statistically significant improvement in 6MWD or mPAP. The safety findings were characterized by higher risks of anemia, peripheral edema, and treatment discontinuation. These results should be interpreted in the context of differences in PH phenotype, dose, comparator, study design, and outcome measurement.

## INTRODUCTION

Pulmonary hypertension (PH) is characterized by elevated pulmonary arterial pressure and may arise from several distinct cardiopulmonary and systemic conditions. Endothelin receptor antagonists (ERAs), including bosentan, ambrisentan, and macitentan are established treatment options for PAH. Macitentan is a second-generation dual ERA with a 50-fold stronger affinity to endothelin A receptors than endothelin B receptors. It has enhanced pharmacologic activity in PAH when compared to bosentan and ambrisentan, and has slower receptor dissociation kinetics.

The SERAPHIN trial showed that a 10mg dose of macitentan reduced the risk of morbidity or mortality events by 45% over the treatment time, establishing macitentan as an important therapeutic option in PAH. Macitentan has subsequently been evaluated in other forms of portopulmonary hypertension, Eisenmenger syndrome, chronic thromboembolic pulmonary hypertension (CTEPH), PH associated with left-heart disease, pediatric PAH, and real-world PAH populations. However, results across these populations have been heterogeneous, and the efficacy and safety of macitentan may differ according to the underlying PH phenotype.

Although previous systematic reviews and meta-analyses have examined the use of macitentan in pulmonary hypertension, additional studies have been published, which makes it possible to better evaluate the safety and efficacy of macitentan, including its long-term effects. This systematic review and meta-analysis aimed to evaluate the efficacy and safety of macitentan compared with control across patients with pulmonary hypertension, with subgroup analyses according to PH classification, including Group 1 PAH, CTEPH (Group 4), and PH associated with left-heart disease (Group 2), to assess whether treatment effects differed across these clinically distinct populations.

## METHODOLOGY

### Study Design

A systematic Review and Meta analysis was performed and reported according to the Preferred Reporting Items for Systematic Reviews and Meta-Analyses (PRISMA 2020).

### Eligibility Criteria Inclusion Criteria

Eligible studies included patients with PH, irrespective of etiology, who received macitentan as monotherapy or in combination with other PH-directed therapy. Eligible comparators were placebo or a concurrent control condition, including standard therapy, non-macitentan treatment, or background therapy without macitentan. Randomized controlled trials, non-randomized interventional studies, and retrospective observational cohort studies were eligible.

### Exclusion Criteria

Studies were excluded if they enrolled participants without pulmonary hypertension; did not include a concurrent comparator; were single-arm studies, case reports, case series, reviews, editorials, letters, comments, protocols, or animal/in vitro studies; did not provide extractable outcome data; could not be retrieved in full text; or represented duplicate/overlapping datasets for which a more complete publication was available.

### Search Strategy

3 databases – PubMed, EMBASE and Cochrane clinical trials – were searched from databse inception to June 2026. The strategy was based on PICO, Medical Subject Headings (MeSH) terms, free-text words. The key works included “pulmonary hypertension,” “pulmonary arterial hypertension,” “pulmonary artery hypertension,” OR “PAH” OR “hypertension, pulmonary,” “macitentan,” “opsumit”. MeSH terms were used when appropriate.

### Screening

Records retrieved from the databases were imported into Rayyan and deduplicated. Two out of six reviewers independently screened titles and abstracts, followed by full-text assessment of potentially eligible studies. Disagreements were resolved by discussion and, when necessary, consultation with a third reviewer. The study-selection process is summarized in a PRISMA 2020 flow diagram (Figure 1).

**Figure 1:**
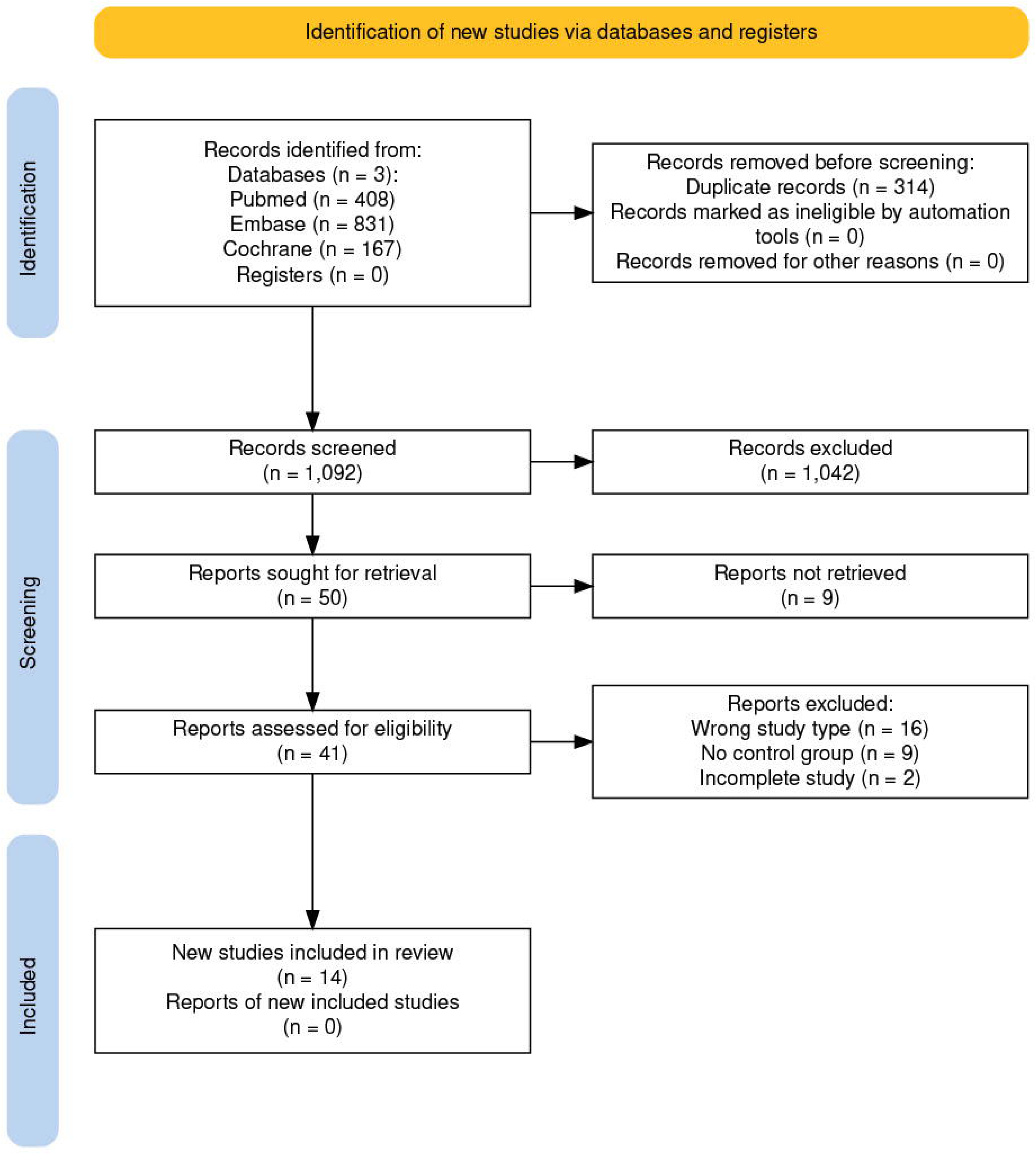
PRISMA Chart

### Data Extraction and Statistical Analysis

A standardized extraction form was used to collect study characteristics and outcome data. Extracted variables included first author, publication year, study design, population and PH subtype, sample size, age, sex, WHO or NYHA functional class, macitentan regimen, comparator, follow-up duration, efficacy outcomes, and adverse events. Mean ± SD was prioritized. When mean and SD were unavailable, medians, interquartile ranges, ranges, confidence intervals, or standard errors of the mean were recorded for subsequent conversion where appropriate. Both follow-up values and changes from baseline were extracted when available. When both were reported, change-from-baseline estimates were prioritized. Alternative summary statistics were converted to mean and SD using the specified conversion approach before meta-analysis.

### Outcomes

Efficacy outcomes were 6MWD, PVR, mPAP, and cardiac index. Safety outcomes were mortality, anemia, peripheral edema, PAH-related hospitalization, dyspnea, pneumonia, headache, discontinuation due to adverse events, and serious adverse events.

### Risk of bias assessment

Risk of bias was assessed using the revised Cochrane Risk of Bias 2 tool for randomized trials and ROBINS-I for non-randomized studies of interventions (Figure 3).

### Statistical Analysis

Random-effects model was used. Continuous outcomes were synthesized using mean differences (MDs) when the same measurement scale was used. Dichotomous outcomes were synthesized using risk ratios (RRs) with 95% confidence intervals (CIs). For outcomes reported as log(RR), pooled estimates and their confidence intervals were exponentiated to obtain RRs and corresponding 95% CIs. Statistical heterogeneity was evaluated using Cochran’s Q statistic, I^2^, and τ^2^. Heterogeneity was interpreted alongside clinical and methodological differences between studies.

Subgroup analyses were performed according to pulmonary hypertension phenotype/World Health Organization (WHO) group where at least two studies contributed data to a given subgroup. Pooled treatment effects were compared across subgroups to explore potential differences in treatment effects between pulmonary hypertension groups. Given the limited number of studies contributing to several outcomes and subgroups, subgroup findings were considered exploratory and interpreted in the context of clinical and methodological heterogeneity.

A two-sided p value <0.05 was considered statistically significant.

Influence analyses were performed to identify studies that contributed substantially to between-study heterogeneity or materially affected the pooled estimates. Funnel plots were examined visually when sufficient studies were available; given the limited number of studies contributing to individual outcomes, assessment of small-study effects and funnel-plot asymmetry was considered exploratory rather than definitive.

All statistical tests were two-sided, and a P value <0.05 was considered statistically significant. Analyses were performed using JASP.

## RESULTS

### Study Selection and characteristics

The database search identified 1,406 records. After removal of 314 duplicates, 1,092 records underwent title and abstract screening. Fifty reports were sought for retrieval, of which 41 were assessed in full text. Twenty-seven reports were excluded at the full-text stage.Fourteen studies comprising 5,198 participants were included in the quantitative synthesis (Figure 1). Eight studies evaluated Group 1 PAH (n = 3,083), three evaluated Group 2 PH-LHD (n = 191), and three evaluated Group 4 CTEPH (n = 425). The included studies comprised randomized controlled trials and observational comparative cohorts and evaluated several PH phenotypes. Study characteristics are summarized in Table 1. Control interventions included placebo, standard of care, tadalafil, bosentan, and other background therapy. Macitentan regimens varied across studies, including 10 mg once daily, weight-based pediatric dosing, combination therapy, and a dose-escalation regimen reaching 75 mg once daily.

**Table 1:** Study Characteristics

| Study | Study design | Follow-up | Population / PH subtype | Total N | Male sex, n (%) | Functional class | Macitentan regimen | Comparator |
| --- | --- | --- | --- | --- | --- | --- | --- | --- |
| TOMORROW 2026 | Prospective, multicenter, open-label, randomized, controlled, parallel-group phase III trial | 12 weeks | Idiopathic, heritable, or PAH associated with congenital heart disease, connective tissue disease, HIV infection, or drug/toxin exposure | 148 | 88 (59.5) | WHO FC I–III | Weight-based:<br>3.5 mg/day (10–<15 kg);<br>5 mg/day (15–<25 kg);<br>7.5 mg/day (25–<50 kg);<br>10 mg/day (≥50 kg) | Standard of care |
| MACITEPH 2025 | Prospective, randomized, double-blind, placebo-controlled, parallel-group, adaptive phase III trial with open-label extension | 28 weeks | Inoperable CTEPH or persistent/recurrent CTEPH after BPA or PEA | 127 | 48 (37.8) | WHO FC II–IV | 10 mg once daily for 4 weeks, followed by 37.5 mg once daily for 4 weeks, then 75 mg once daily | Placebo |
| MERIT 2024 | Multicenter, randomized, double-blind, placebo-controlled, parallel-group phase II trial | 24 weeks | Inoperable CTEPH | 80 | 29 (36) | WHO FC II–IV | 10 mg once daily | Placebo |
| SOPRANO 2024 | Prospective, multicenter, randomized, double-blind, placebo-controlled, parallel-group phase II trial | 12 weeks | Persistent PH following LVAD implantation | 57 | 45 (78.9) | WHO FC II–III | 10 mg once daily | Placebo |
| MELODY 2018 | Multicenter, randomized, double-blind, placebo-controlled phase II trial | 12 weeks | Combined post- and precapillary PH due to left heart disease (CpcPH) | 63 | 22 (34.9) | NYHA FC II–III | 10 mg once daily | Placebo |
| Thor et al., 2019 | Single-center retrospective cohort study | 2 years | Inoperable or residual CTEPH | 112 | 47 (42.0) | WHO FC I–IV | 10 mg once daily | Bosentan |
| Tynan et al., 2019 | Single-center retrospective cohort study | 18 months | Confirmed PAH | 37 | 7 (19.3) | NR | 10 mg once daily | Stable sildenafil and/or bosentan/ambri-sentan therapy |
| Grunig et al., 2024 | Prospective, multicenter, double-blind, randomized, active-controlled, triple-dummy, parallel-group, adaptive phase III trial | NR | Idiopathic, heritable, drug/toxin-induced, or PAH associated with connective tissue disease, HIV infection, portal hypertension, or corrected congenital heart disease | 79 | 16 (20.3) | WHO FC II–III | Macitentan 10 mg once daily + tadalafil 40 mg once daily | Tadalafil monotherapy |
| Gatzoulis et al.,<br>2019 | Multicenter,<br>double-blind,<br>randomized,<br>placebo-controlled<br>phase III trial<br>(MAESTRO) | 16 weeks | Eisenmenger<br>syndrome | 226 | 76 (33.6) | WHO FC<br>II–III | 10 mg once<br>daily | Placebo |
| Krowka et al.,<br>2018 | Prospective,<br>multicenter,<br>randomized,<br>double-blind,<br>placebo-controlled<br>phase IV trial | 12 weeks | Portopulmonary<br>hypertension<br>(PoPH) | 85 | 42 (49.4) | NR | 10 mg once<br>daily | Placebo |
| Pulido et al., 2013 | Multicenter,<br>double-blind,<br>placebo-controlled,<br>event-driven phase<br>III trial | 2.5 years | Symptomatic<br>WHO Group 1<br>PAH,<br>idiopathic/heritabl<br>e or associated<br>PAH | 742 | 177 (23.9) | WHO FC<br>II–IV | 3 or 10 mg<br>once daily | Placebo |
| Shah et al., 2025 | Randomized,<br>double-blind,<br>multicenter trial | 24 weeks | HF with LVEF<br>≥40% and<br>evidence of LV<br>hypertrophy or LA<br>enlargement,<br>elevated natriuretic<br>peptides, and<br>evidence of PVD | 142 | 55 (38.7) | NYHA FC<br>II–III | 10 mg once<br>daily | Placebo |
| Kashaki et al.,<br>2025 | Randomized,<br>double-blind,<br>non-inferiority,<br>parallel-group<br>clinical trial | 12 days | Newborns >72 h<br>with suspected<br>PPHN and<br>persistent<br>respiratory distress<br>and/or cyanosis<br>despite supportive<br>management | 60 | 29 (48.3) | NR | Macitentan 1<br>mg/kg/dose<br>every 12 h +<br>oral sildenafil<br>every 12 h | Bosentan +<br>oral sildenafil |
| Benza et al., 2020 | Retrospective cohort<br>study using<br>Medicare claims<br>data | 21–28<br>months | PAH identified<br>using ICD-9/10<br>codes | 3,240 | 851 (26.26) | NR | 10 mg once<br>daily | Ambrisentan<br>and bosentan |

**Abbreviations:** BPA, balloon pulmonary angioplasty; CpcPH, combined post- and precapillary pulmonary hypertension; CTEPH, chronic thromboembolic pulmonary hypertension; FC, functional class; HF, heart failure; HIV, human immunodeficiency virus; ICD, International Classification of Diseases; LA, left atrial; LV, left ventricular; LVAD, left ventricular assist device; LVEF, left ventricular ejection fraction; MAESTRO, Macitentan in Eisenmenger Syndrome Treatment Randomized, Controlled, Multicenter Study; NR, not reported; PAH, pulmonary arterial hypertension; PAWP, pulmonary artery wedge pressure; PEA, pulmonary endarterectomy; PH, pulmonary hypertension; PPHN, persistent pulmonary hypertension of the newborn; PoPH, portopulmonary hypertension; PVD, peripheral vascular disease; SoC, standard of care; WHO, World Health Organization.

**Table 2:**
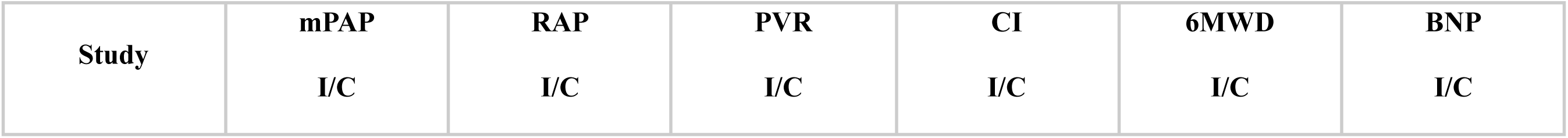
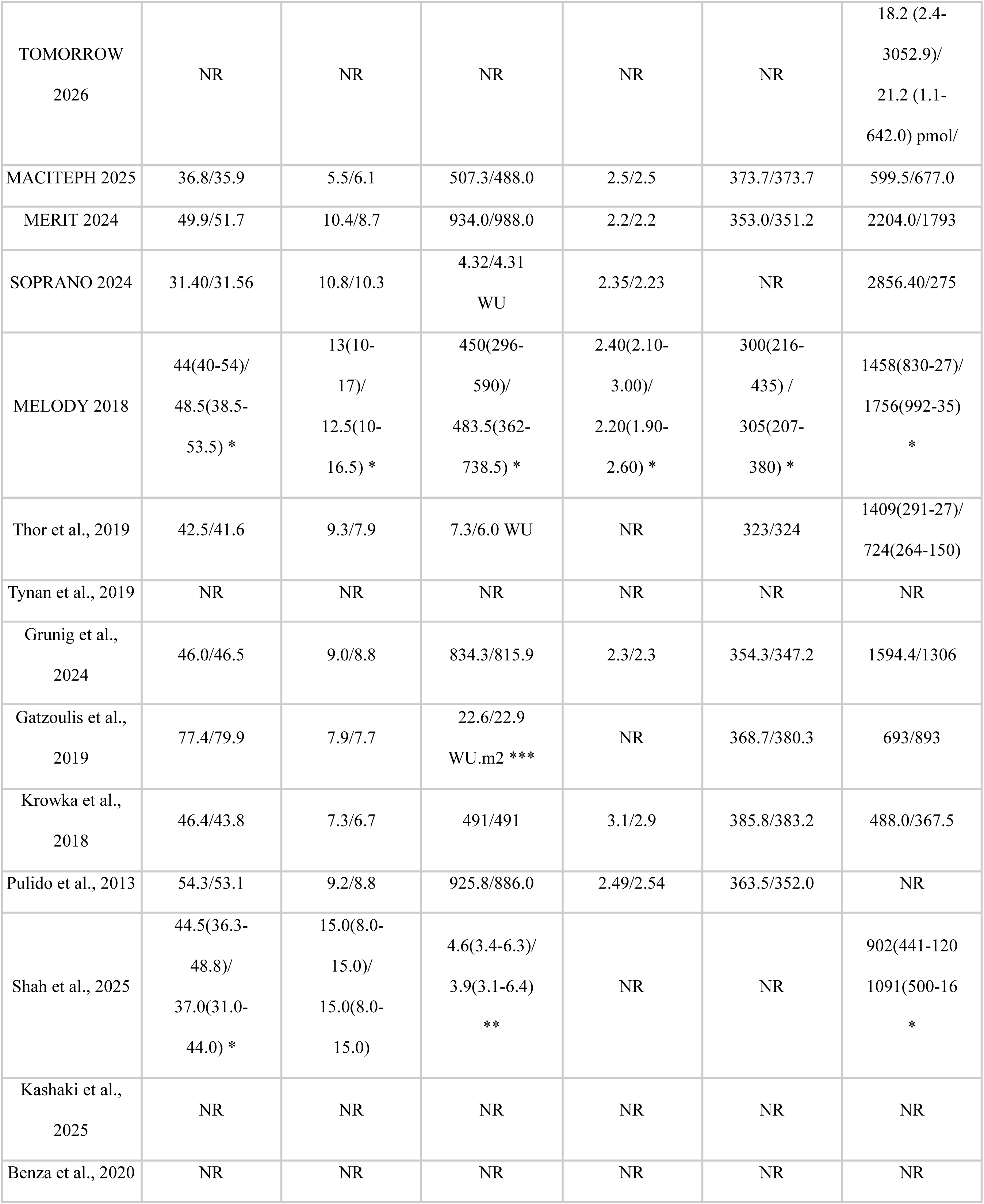
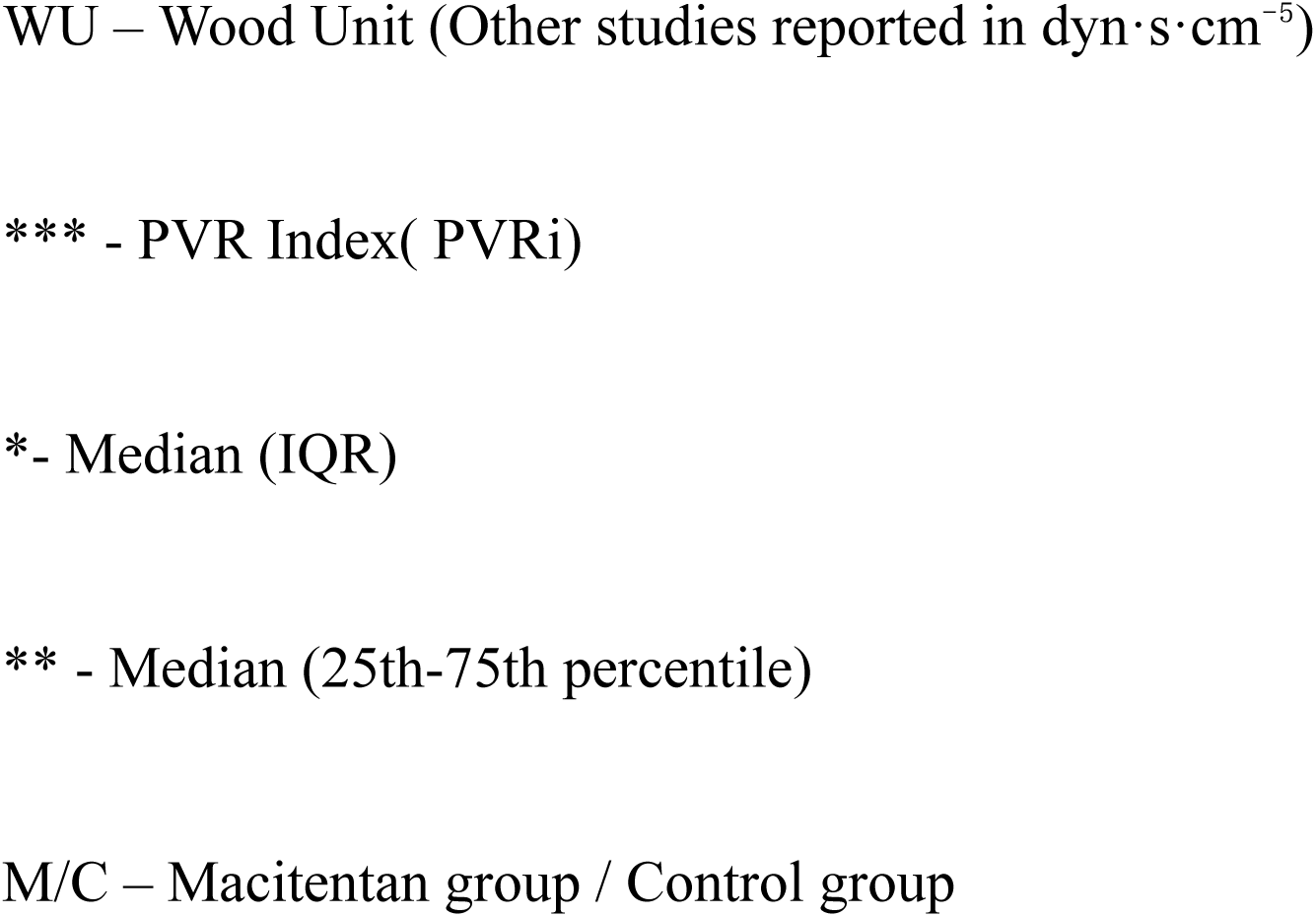
Baseline Hemodynamic Characteristics of Patients in Included Studies.

### Efficacy Outcomes

#### 6-Minute-Walking-Distance (6MWD)

Six studies evaluated change in 6MWD. Macitentan was not associated with a statistically significant improvement in 6MWD (MD, 5.04m; 95% CI, -15.06 to 25.14; p = 0.547).

Subgroup analyses according to PH classification were performed where at least two studies contributed to a subgroup. Three studies contributed to the CTEPH subgroup, in which the pooled effect was 7.13 m (95% CI, −57.27 to 71.53; P = 0.681), with substantial heterogeneity (I^2^ = 72.81%). Two studies contributed to the PAH subgroup, with a pooled effect of 9.71 m (95% CI, −121.59 to 141.00; P = 0.520) and moderate-to-substantial heterogeneity (I^2^ = 66.42%). Only 1 study contributed to the PH-LHD subgroup, and therefore a pooled estimate could not be calculated for this group. There was no statistically significant difference in treatment effects between the CTEPH and PAH subgroups (Q(1) = 0.02; P = 0.887).

An RCT-only sensitivity analysis excluding the single observational study yielded a pooled MD of 4.60 m (95% CI, −20.93 to 30.13; I^2^ = 73%), which was consistent with the primary analysis.

#### Pulmonary Vascular Resistance (PVR)

Six studies also evaluated pulmonary vascular resistance, which was significantly reduced with macitentan in comparison to the control group (MD, -134.50 dyn·s·cm⁻⁵; 95% CI, -226.1 to -42.86; p = 0.013).

Subgroup analyses according to PH classification were performed because at least two studies contributed to each subgroup. Two studies contributed to the CTEPH subgroup, with a pooled MD of −182.10 dyn·s·cm⁻⁵ (95% CI, −917.70 to 553.50). Two studies contributed to the PH-LHD subgroup, with a pooled MD of −62.75 dyn·s·cm⁻⁵ (95% CI, −153.90 to 28.44). Two studies contributed to the PAH subgroup, with a pooled MD of −212.60 dyn·s·cm⁻⁵ (95% CI, −1180.00 to 754.40). Within-subgroup heterogeneity was not statistically significant for CTEPH (Q(1) = 0.85; P = 0.421), PH-LHD (Q(1) = 0.09; P = 0.768), or PAH (Q(1) = 1.94; P = 0.163).

There was a statistically significant difference in pooled treatment effects across the three PH classification subgroups (Q(2) = 7.93; P = 0.019). However, the confidence intervals for all three subgroup estimates crossed the null, and the subgroup analysis was considered exploratory given the small number of studies contributing to each subgroup.

#### Mean Pulmonary Arterial Pressure (mPAP)

Five studies assessed mean pulmonary arterial pressure (mPAP). Macitentan was not associated with a statistically significant reduction in mPAP compared with control (MD, −2.70 mmHg; 95% CI, −6.46 to 1.05; P = 0.116). Moderate heterogeneity was observed (I^2^ = 66.1%).

Subgroup analyses according to PH classification were performed where at least two studies contributed to a subgroup. Two studies contributed to the CTEPH subgroup, with a pooled MD of −2.84 mmHg (95% CI, −14.14 to 8.45; P = 0.193). Two studies contributed to the PH-LHD subgroup, with a pooled MD of 0.16 mmHg (95% CI, −1.62 to 1.94; P = 0.465). No pooled estimate was calculated for the PAH subgroup because fewer than two studies contributed data. Within-subgroup heterogeneity was not statistically significant for either CTEPH (Q(1) = 0.47; P = 0.491) or PH-LHD (Q(1) = 0.01; P = 0.928). There was a statistically significant difference between the CTEPH and PH-LHD subgroup estimates (Q(1) = 11.09; P < 0.001). This subgroup finding was considered exploratory given the small number of contributing studies.

#### Cardiac Index (CI)

Six studies evaluated cardiac index. Macitentan was associated with a statistically significant increase in cardiac index (MD, 0.44 L/min/m2; 95% CI, 0.22 to 0.67; p = 0.004).

Subgroup analyses according to PH classification were performed where at least two studies contributed to each subgroup. Two studies contributed to the CTEPH subgroup, with a pooled MD of 0.53 L/min/m^2^ (95% CI, −0.04 to 1.09; P = 0.054). Two studies contributed to the PH-LHD subgroup, with a pooled MD of 0.21 L/min/m^2^ (95% CI, −2.07 to 2.50; P = 0.446). Two studies contributed to the PAH subgroup, with a pooled MD of 0.55 L/min/m^2^ (95% CI, −0.34 to 1.43; P = 0.080). There was no statistically significant difference in treatment effects across the three PH classification subgroups (Q(2) = 3.08; P = 0.215).

Heterogeneity estimates in the working analysis were I^2^=65.6%, 58.2%, 66.1%, and 61.5% for 6MWD, PVR, mPAP, and cardiac index, respectively. Influence diagnostics identified MACiTEPH for 6MWD, SOPRANO for PVR and cardiac index, and Krowka et al. for mPAP as influential studies; SOPRANO for cardiac index and Krowka et al. for mPAP were also identified as outliers. The corresponding forest plots are shown in Figure 2.

**Figure 2:**
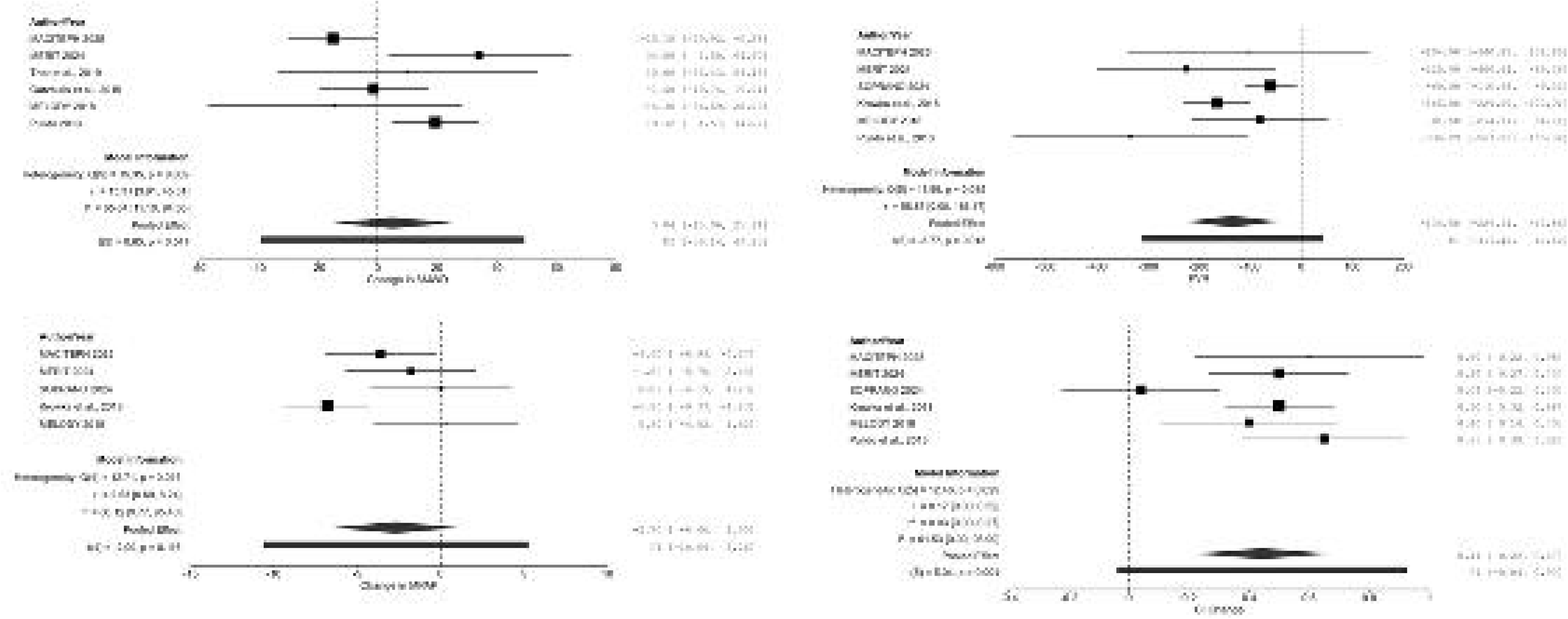
Forest plots of the effects of macitentan compared with placebo on functional and hemodynamic outcomes. The figure presents pooled effects for (A) change in 6-minute walk distance (6MWD), (B) pulmonary vascular resistance (PVR), (C) change in mean pulmonary arterial pressure (mPAP), and (D) change in cardiac index (CI).

**Figure 3:**
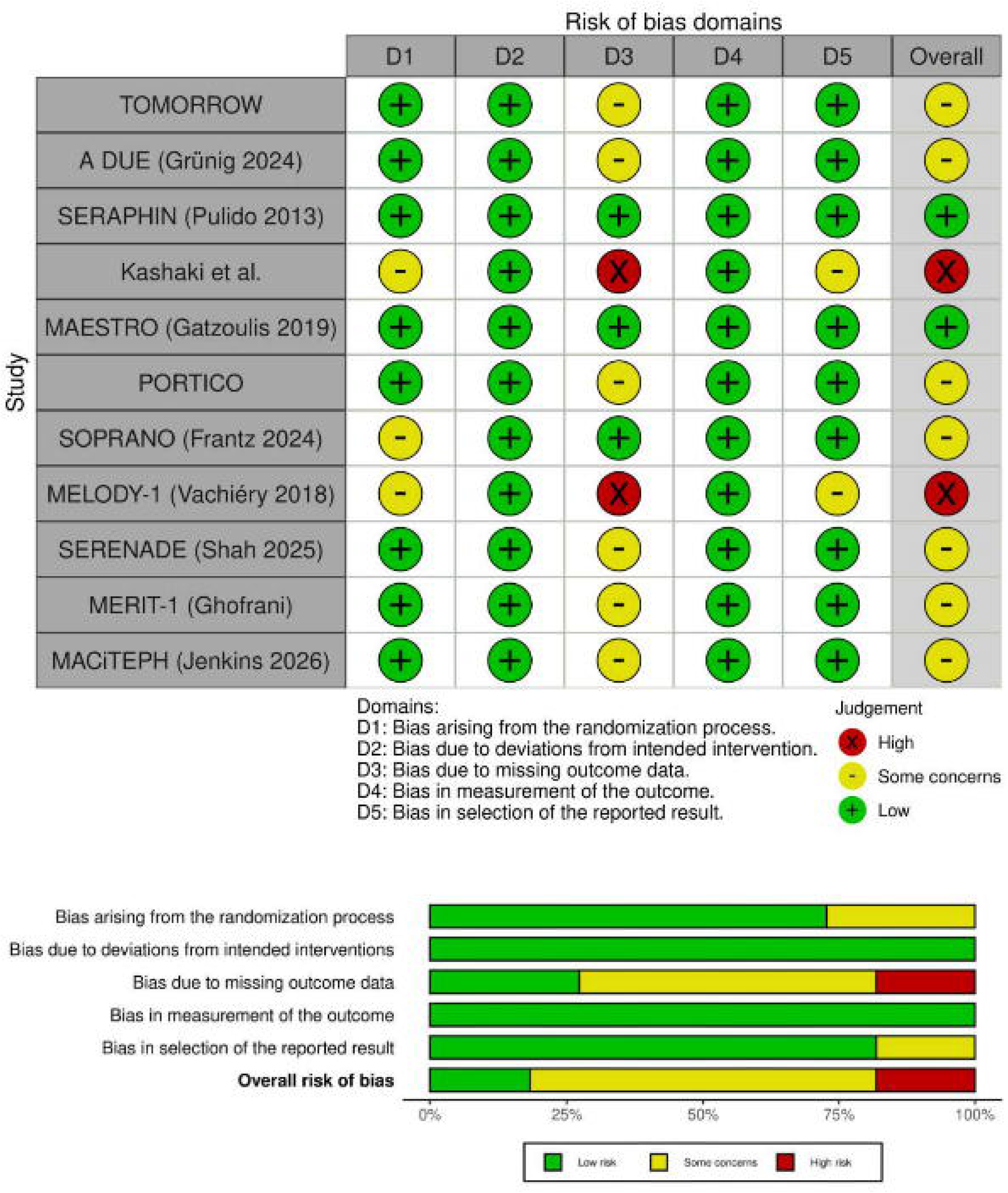
ROB Assessment for Included RCTs

**Figure 4:**
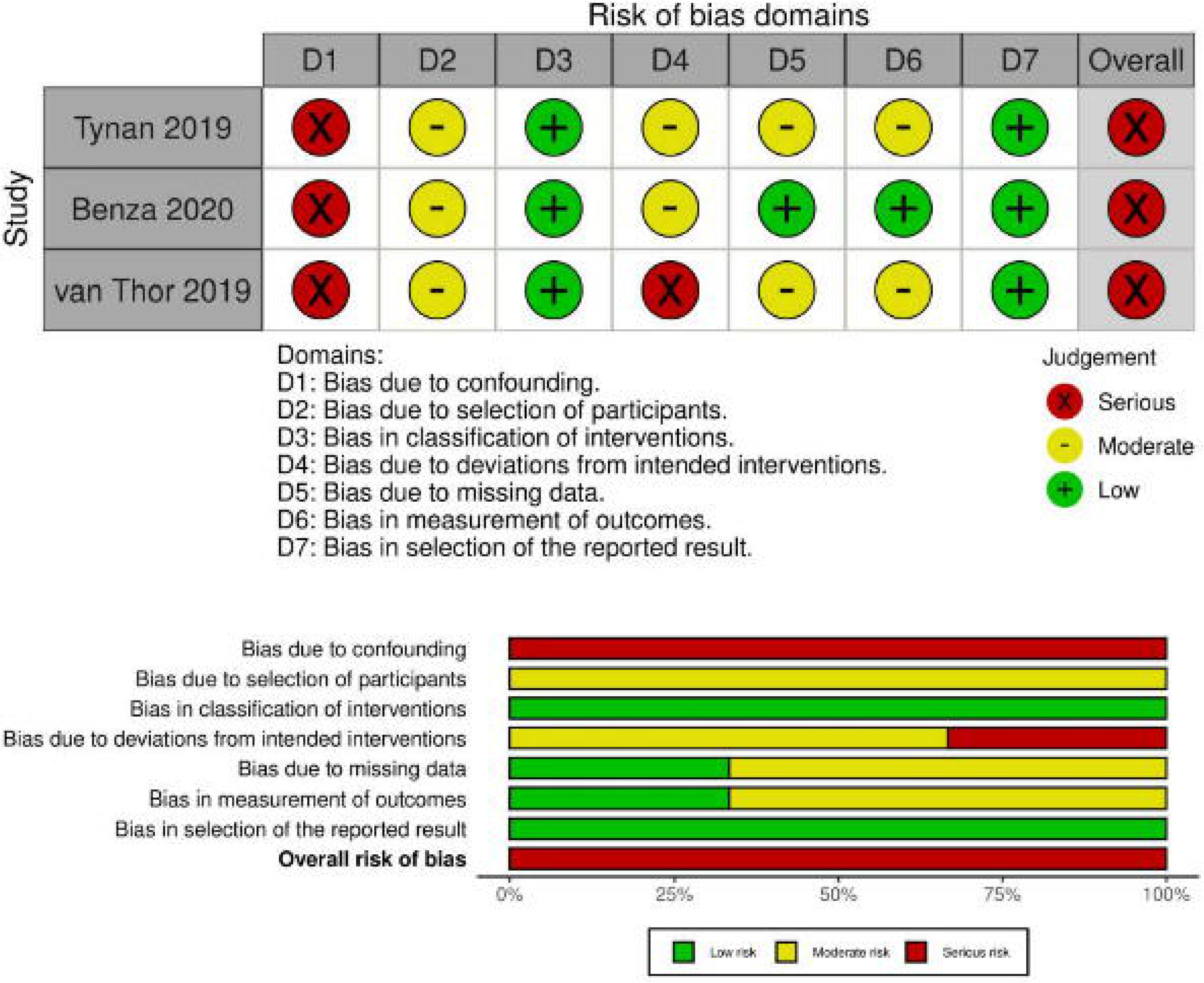
ROB Assessment for Included Observational Studies

### Safety Outcomes

#### Mortality

A total of 14 studies evaluated overall mortality, and macitentan demonstrated a statistically significant reduction in mortality compared with the control group (pooled RR 0.79; 95% CI, 0.73 to 0.86; p < 0.001).

#### Anemia

Twelve studies evaluated the incidence of anemia, showing a statistically significant higher risk in the macitentan group (RR, 2.97; 95% CI, 1.86 to 4.71; p < 0.001).

#### Peripheral Edema

Ten studies evaluated peripheral edema, which revealed a statistically significant increase in risk with macitentan (RR, 1.41; 95% CI, 1.001 to 2.00; p =0.050).

#### Discontinuation due to adverse events

Twelve studies assessed discontinuation due to adverse events, demonstrating a statistically significant increase in treatment discontinuation in the macitentan group compared to control (RR, 1.90; 95% CI, 1.04 to 3.46; p = 0.038).

#### Other safety outcomes

In contrast, no statistically significant differences were observed between macitentan and control groups for PAH-related hospitalization across six studies (RR, 0.93; 95% CI, 0.55 to 1.60; p = 0.767), dyspnea across five studies (RR, 1.15, 95% CI, 0.76 to 1.73; p = 0.400), pneumonia across four studies (RR, 1.54; 95% CI, 0.21 to 11.42; p = 0.540), headache across six studies (RR, 1.31; 95% CI, 0.9 to 1.92; p = 0.130), or serious adverse events (SAEs) across ten studies (RR, 1.26; 95% CI, 0.93 to 1.7; p = 0.113).

Heterogeneity was low for most safety outcomes, with no significant heterogeneity detected for mortality, anemia, PAH-related hospitalization, dyspnea, headache, peripheral edema, or discontinuation due to adverse events. Low-to-moderate heterogeneity was observed for pneumonia (I^2^ = 36.1%) and serious adverse events.

Because fewer than 10 studies contributed to each safety outcome, funnel-plot asymmetry findings were interpreted cautiously. Influence analyses identified Pulido et al. as influential across several safety endpoints, with additional influence attributed to Paoli et al. for hospitalization and Benza et al. and Shah et al. for mortality

### Sensitivity analyses

A sensitivity analysis restricted to randomized controlled trials was performed for 6MWD because one of the six contributing studies was observational. Five randomized trials contributed to the sensitivity analysis, yielding an MD of 4.60 m (95% CI, −20.93 to 30.13; I^2^=73%). The primary analyses of PVR, mPAP, and cardiac index included only randomized controlled trials; therefore, no additional randomized-trial-only sensitivity analysis was required for these outcomes. Because all studies contributing to PVR, mPAP, and cardiac index were placebo-controlled randomized trials, separate placebo-controlled sensitivity analyses were also not required for these outcomes.

## Discussion

The present systematic review and meta-analysis synthesized 14 studies involving 5,198 participants with WHO Group 1 pulmonary arterial hypertension (PAH), Group 2 PH associated with left-heart disease (PH-LHD), and Group 4 chronic thromboembolic pulmonary hypertension (CTEPH). We found favorable pooled effects for PVR and cardiac index, whereas effects on 6MWD and mPAP were not statistically significant. Safety analyses showed higher risks of anemia, peripheral edema, and discontinuation due to adverse events. These findings highlight a consistent hemodynamic effect of macitentan across several PH populations, while functional outcomes appear to depend more strongly on disease phenotype, treatment context, and the relationship between pulmonary vascular improvement and overall exercise capacity.

The significant reduction in PVR is an important finding because PVR represents a major component of right-ventricular (RV) afterload and is closely related to progression of pulmonary vascular disease.

Individual trials have reported favorable hemodynamic effects with macitentan in PAH and CTEPH, including PORTICO and MERIT-1. Despite this convergence at the level of individual trials, the phenotype-specific pooled estimates in the present analysis were imprecise because only two studies contributed to each subgroup, the overall direction and magnitude of the pooled effect support a clinically relevant pulmonary vascular response to macitentan. The smaller effect observed in PH-LHD is also consistent with the distinct pathophysiology of pulmonary hypertension arising from left-heart disease rather than primary pulmonary vascular disease.

Macitentan was also associated with a significant increase in cardiac index (0.44 L/min/m^2^). This provides an added hemodynamic signal of improved cardiopulmonary performance as reducing PVR lowers RV afterload and can improve RV-pulmonary arterial coupling and forward flow. The direction of effect was consistent across the CTEPH, PH-LHD, and PAH subgroups, with increases of 0.53, 0.21, and 0.55 L/min/m^2^, respectively, and there was no significant difference between subgroups (Q(2) = 3.08; P = 0.215). The consistency across phenotypes suggests that improvement in cardiac output may represent a common downstream consequence of reduced pulmonary vascular load. However, a pooled increase in cardiac index should not automatically be interpreted as evidence of improved functional status. The absence of a statistically significant effect on 6MWD illustrates the distinction between hemodynamic and patient-centered outcomes. The latter may be influenced by exercise limitation, comorbidity, right-to-left shunting, chronic hypoxemia, and other factors that are not captured by changes in pulmonary vascular resistance alone.

The absence of a significant reduction in mPAP provides an important contrast to the PVR and cardiac index findings. Mean pulmonary arterial pressure is determined by the interaction between pulmonary vascular resistance, cardiac output, and left atrial filling pressure. Consequently, a reduction in PVR accompanied by a change in cardiac output does not necessarily produce a proportionate reduction in mPAP. Krowka et al. was identified as an outlier for mPAP possibly because its population – patients with portopulmonary hypertension – often carry a component of hyperdynamic, high cardiac output circulation from their liver disease that can produce mPAP behavior distinct from that seen in classic pre-capillary PAH. Moreover, the stronger reduction in CTEPH compared to PH-LHD in subgroup analysis is consistent with the favorable pulmonary vascular effects observed in MERIT-1, whereas the absence of a comparable effect in PH-LHD reflects the fundamentally different hemodynamic drivers of pulmonary hypertension in left-heart disease. These findings reinforce that changes in pulmonary pressure should be interpreted alongside PVR and cardiac output rather than as an isolated marker of treatment response.

In contrast, macitentan did not significantly improve 6MWD. MACiTEPH , a phase 3 trial using a 75 mg dose of macitentan, three times the standard 10 mg used in most other included studies was stopped early for futility after a planned interim analysis showed no separation from placebo on its primary 6MWD endpoint, and it was identified here as an influential study for the 6MWD outcome. One possible explanation is dose-related pharmacological heterogeneity. At the standard dose, macitentan produces substantial ETA receptor blockade while retaining partial ETB receptor activity; greater ETB blockade at higher exposure could theoretically attenuate some of the vasodilatory effects associated with preserved ETB signaling. Although this mechanism was not directly evaluated in MACiTEPH, it provides a plausible pharmacological explanation for the divergent functional findings at different doses.

MAESTRO, a dedicated trial in patients with Eisenmenger syndrome, similarly found no significant improvement in exercise capacity despite favorable changes in NT-proBNP and hemodynamic measures. Chronic hypoxemia, fixed cardiopulmonary limitations, and long-standing right-to-left shunting may limit the extent to which improvements in pulmonary vascular tone translate into measurable gains in walking distance in this population. Thus, the absence of a significant pooled 6MWD benefit does not necessarily contradict the observed hemodynamic improvements. The modest functional improvement reported in adult PAH populations such as SERAPHIN, compared with the lack of significant benefit in MACiTEPH and MAESTRO, further suggests that dose and underlying phenotype influence the extent to which pulmonary vascular improvement translates into measurable functional gain.

Macitentan was associated with a 21% relative reduction in overall mortality (RR 0.79; 95% CI 0.73–0.86). This finding is consistent with the broader clinical evidence for macitentan in PAH, although SERAPHIN itself evaluated a composite morbidity/mortality endpoint rather than mortality alone. Because SERAPHIN was substantially larger than many of the other included studies, it carries considerable weight in pooled estimates. Shah et al. (SERENADE), which enrolled patients with Group 2 PH associated with HFpEF/HFmrEF, was also identified as influential in the mortality analysis and reported no clinical benefit from macitentan in that population. The contrast between these populations suggests that the mortality signal observed in the overall analysis is driven predominantly by the established Group 1 PAH evidence and should not automatically be extrapolated to Group 2 PH-LHD.

These results have several clinical implications that follow phenotype. Within Group 1 PAH, the included evidence spans idiopathic and connective-tissue-associated PAH, portopulmonary hypertension, Eisenmenger syndrome, and pediatric disease.[1,2,6,7] Across this spectrum, the hemodynamic effects of macitentan were more consistent than the effects on functional capacity.

Group 2 PH-LHD represents a similarly heterogeneous treatment population. SOPRANO enrolled patients with continuous-flow left ventricular assist devices, whereas MELODY-1 and SERENADE evaluated patients with PH associated with underlying left-heart disease without LVAD support.[9-11] The mechanisms contributing to pulmonary hypertension in these populations may differ substantially, as may the effects of background cardiac therapies and mechanical circulatory support. These differences provide important context for the variable response to macitentan observed across Group 2 studies.

In Group 4 CTEPH, MERIT-1 reported favorable findings with macitentan 10 mg, whereas MACiTEPH evaluated a substantially higher 75-mg dose and was stopped early for futility.[3,5] The contrast between these studies raises the possibility that dose, treatment duration, patient selection, and endpoint characteristics may influence the observed efficacy of macitentan in CTEPH. The results also emphasize the importance of distal pulmonary microvasculopathy as a potential therapeutic target in CTEPH alongside the mechanical component of chronic thromboembolic obstruction.

The safety findings are broadly consistent with known adverse effects associated with ERA therapy, particularly reductions in hemoglobin and peripheral edema. However, they should be interpreted in the context of a mixed evidence base. Randomized trials and observational comparative cohorts differed in design, population, comparator, follow-up, and potential sources of bias.

From a practical standpoint, these findings support an individualized approach to both prescribing and monitoring. Baseline and periodic hemoglobin assessment is reasonable given the magnitude of the anemia , particularly in patients who are already anemic or who have limited physiological reserve.

Volume status should be assessed clinically rather than assumed to be benign, since peripheral edema can reflect either an expected drug effect or early right heart decompensation, and the two require different responses. Perhaps most importantly, clinicians and patients should be counseled that a hemodynamic improvement on macitentan does not guarantee a matching improvement in walking distance or symptoms, particularly outside typical adult idiopathic PAH, a distinction that matters for how treatment response is framed and assessed in clinic.

A major strength of this analysis is that, although the overall evidence base included multiple study designs and comparator types, the studies contributing to the pooled efficacy outcomes were placebo-controlled randomized controlled trials (except for 6MWD for which sensitivity analysis was performed). Consequently, the efficacy estimates were not influenced by the methodological heterogeneity introduced by observational studies, active-comparator designs, or nonrandomized evidence. The principal source of heterogeneity in the efficacy analyses was therefore clinical, particularly variation in PH phenotype, disease characteristics, treatment context, and study populations. This provides greater internal consistency for interpretation of the efficacy findings while allowing the broader evidence base to contribute to safety and real-world outcomes.

The main limitations are clinical heterogeneity across PH phenotypes and small numbers of studies within individual subgroups, which limited phenotype-specific estimates. Heterogeneity was particularly evident for 6MWD, while influence analyses identified several studies with substantial effects on individual outcomes. The safety analyses were more methodologically heterogeneous because they included both randomized and observational studies with different comparators and follow-up periods. Finally, fewer than ten studies contributed to individual outcomes, limiting assessment of publication bias

Future research should prioritize dose ranging comparisons of macitentan 10 mg versus 75 mg to directly test whether the ETB-mediated mechanism proposed here for the MACiTEPHcontributes to differences in functional efficacy. Phenotype specific trials that distinguish device related from primary myocardial

PH-LHD rather than pooling both under Group 2, and standardized 6MWD protocols across future PAH and CTEPH trials to reduce the substantial residual heterogeneity observed in this analysis should be done. Individual patient data meta-analysis could also determine whether baseline PVR, RV dysfunction, PH phenotype, or background therapy modifies response to macitentan.

In summary, macitentan produces a consistent and meaningful improvement in pulmonary vascular resistance and cardiac index, together with a mortality benefit that appears to be driven predominantly by Group 1 PAH evidence, while functional and pressure-based outcomes are shaped by dose, phenotype, and population factors that this analysis was able to identify at the level of individual contributing trials even where the pooled estimates alone could not. The overall clinical message is that macitentan’s benefit to risk profile should be assessed phenotype by phenotype, informed by the specific trials that make up the evidence for that phenotype, rather than treated as a single uniform effect across all forms of pulmonary hypertension.

## Conclusion

Macitentan significantly improved PVR and cardiac index, with benefits varying across PH phenotypes. Functional and pressure outcomes were less consistent, while mortality benefit was driven predominantly by Group 1 PAH evidence. These findings support phenotype-specific assessment of macitentan efficacy and safety.

## Data Availability

All data produced in the present study are available upon reasonable request to the authors

